# Scaling Sensor Metadata Extraction for Exposure Health Using LLMs

**DOI:** 10.1101/2025.08.21.25334173

**Authors:** Fatemeh Shah-Mohammadi, Sunho Im, Julio C. Facelli, Mollie R. Cummins, Ram Gouripeddi

**Affiliations:** Department of Biomedical Informatics, The University of Utah, Salt Lake City, UT 84108, United States; College of Nursing, The University of Utah, Salt Lake City, UT 84108, United States; Utah Clinical and Translational Science Institute, The University of Utah, Salt Lake City, UT 84108, United States

**Author notes:** Corresponding Author: Fatemeh Shah-Mohammadi, PhD, Department of Biomedical Informatics, School of Medicine, University of Utah, Salt Lake City, Utah, USA.

**Keywords:** Exposure health, Sensor, Metadata, LLM, GPT

## Abstract

**Background:** The rapid evolution and diversity of sensor technologies, coupled with inconsistencies in how sensor metadata is reported across formats and sources, present significant challenges for generating exposomes and exposure health research.

**Objective:** Despite the development of standardized metadata schemas, the process of extracting sensor metadata from unstructured sources remains largely manual and unscalable. To address this bottleneck, we developed and evaluated a large language model (LLM)-based pipeline for automating sensor metadata extraction and harmonization from exposure health literature publicly available.

**Methods:** Using GPT-4 in a zero-shot setting, we constructed a pipeline that parses full-text PDFs to extract metadata and harmonizes output into structured formats. Results: Our automated pipeline achieved substantial efficiency gains in completing extractions much faster than manual review and demonstrated strong performance with average accuracy and precision of 94.74%, recall of 100%, and F1-score of 97.28%.

**Conclusions:** This study demonstrates the feasibility and scalability of leveraging LLMs to automate sensor metadata extraction for exposure health, reducing manual burden while enhancing metadata completeness and consistency. Our findings support the integration of LLM-driven pipelines into exposure health informatics platforms.

## Introduction

Sensors, defined by the NIH as tools that detect, transmit, or report data on biological, chemical, or physical processes, are characterized by metadata describing their principles, accuracy, calibration, and deployment features [9,26,28–32]. The diversity and rapid evolution of sensors [1,2] coupled with the diverse formats and inconsistent structures of sensor metadata create significant challenges for selecting, integrating, and utilizing optimal sensors for exposure health studies. There is a widely recognized need for systematic approaches to improve the harmonization and integration of sensor metadata, for the effective use of emerging technologies in exposure health research [3-7].

Despite this expansion, significant challenges persist in applying sensors and sensor data to exposome research [35]. For prospective sensor use in the quantification of the exposome, these challenges include selecting appropriate sensors, deploying and monitoring sensor networks, managing data streams, pre-processing sensor data and integrating with other data sources for analysis [36,37]. For secondary use of existing sensor-based measurement, including real-world data resources, for the assimilating exposomes, there is a need to find and understand information about sensor characteristics and capabilities for appropriate harmonization and utilization of data for generating exposomes [38,39]. Generally underdeveloped data standards in exposure health, and the specific lack of standardization among sensors and sensor data, further complicate the process of sensor-based exposure health. All of these challenges become more daunting as research questions necessitate multiple and varied sensors, or measurements of varied provenance.

To address this complexity, standardized models are needed to ensure metadata is consistent and computable. The Sensors and Metadata for Analytics and Research in Exposure Health (SMARTER) project developed the Sensor Common Metadata Specifications, a logical model organized into three domains: Instrument, Deployment, and Output [4,8,27]. The Instrument domain captures physical and functional attributes, including capabilities and validation; the Deployment domain describes usage context, including calibration procedures; and the Output domain specifies measured entities and associated metadata. Collectively, these domains provide a comprehensive framework for describing sensor technologies. The SMARTER model is being designed for integration into informatics platforms such as the Exposure Health Informatics Ecosystem (EHIE) [20] to promote interoperability and reuse. In this study, we specifically focus on the “Instrument” domain, and within that, the “Instrument” entity. This entity represents the core metadata about the physical sensor itself, including details necessary to identify, classify, and interpret its use in exposure health contexts. The associated attributes describe technical specifications, operational characteristics, and contextual deployment details.

Currently, researchers must parse sensor documentation, including technical datasheets, manufacturer manuals, exposure health literature, and unstructured web content, to identify relevant sensor metadata. Populating SMARTER schema from heterogeneous, unstructured documentation also remains largely manual and unscalable. As the exposome increasingly rely on diverse sensor technologies, scalable and automated approaches to metadata extraction are essential to support efficient data integration into research workflows [24]. Natural Language Processing (NLP) offers a promising solution to the challenges associated with extracting sensor metadata from unstructured and heterogeneous textual sources. By leveraging techniques such as named entity recognition, relation extraction, and language modeling [11-13], NLP systems can automatically identify, structure and harmonize relevant metadata fields embedded in sensor documentation. Recent advancements in large language models (LLMs) greatly enhance this capability by enabling accurate understanding of complex domain-specific language, even in the absence of rigid templates or standard formats [10,14] or costly training on specifically curated repositories.

NLP, when implemented using LLMs, has shown considerable promise in the automated extraction of metadata across diverse domains. For instance, prior work has demonstrated the capability of LLMs to extract metadata from scholarly research articles, aiming to identify references to datasets and associated metadata descriptors [15]. Digital built environment has leveraged transformer-based language models and custom tokenizers to assign semantic tags to building sensor metadata, demonstrating over 70% tagging accuracy in real-world scenarios [18,19].

Despite these advancements, there is currently no systematic or automated framework dedicated to the extraction of metadata about sensors themselves, in the context of exposure health, from diverse documents describing sensors and sensor deployments. This gap represents a significant opportunity for applying LLM-based NLP methods to automate the extraction and harmonization of human exposure sensor metadata across diverse sources. Our work addresses this gap by automating extraction and harmonization of instrument-level metadata to populate the SMARTER schema and integrate it into the overarching EHIE architecture [33]. The purpose of this study is to develop and evaluate an LLM-based approach for automatically extracting and harmonizing sensor metadata from unstructured exposure health literature. Specifically, we built and assessed a prototypical zero-shot extraction pipeline using GPT-4, iteratively refining prompts and applying NLP post-processing to align outputs with the SMARTER metadata schema [4,8]. We adopted a zero-shot strategy to avoid the substantial cost and time required to curate a task-specific labeled corpus across heterogeneous sources, a process known to be a major bottleneck that typically demands domain experts for annotation [25]. This automated framework addresses a critical bottleneck in environmental health research by significantly reducing the manual burden of metadata curation, improving the quality and consistency of extracted information, and enhancing the scalability of metadata repository development.

## Methods

In this study we focus on the Instrument domain, specifically the Instrument entity which contains the core attributes needed to identify, classify, and interpret a sensor for exposure-health applications. The associated attributes describe technical specifications, operational characteristics, and contextual deployment details. The full list of attributes defined for this entity, along with descriptions can be find in the Supplementary Material. These attributes include both required and optional fields, covering aspects such as model name, manufacturer, measured entities, power source, dimensions, usage context, and maintenance recommendations.

### Data Source

While diverse resources document sensor metadata, such as websites, datasheets, and catalogs, we focused on extracting the metadata embedded in PDF-format from published research articles. Ten papers were randomly selected from a pool of air exposure health sensor studies identified through a prior scoping review of exposure health research studies. The original scoping review focused on identifying and screening studies involving environmental sensors. From the resulting studies involving environmental sensors, a subset of 37 studies specifically related to air quality sensors were found, and 10 were randomly chosen for further analysis. We considered only 10 papers to reduce the manual review time to a manageable time. This phase served as a foundational step for evaluating the feasibility and accuracy of LLM-based sensor metadata extraction.

In this study, we used the base version of the GPT-4o model for generative question answering [46]. This model was accessed via OpenAI’s API in zero-shot learning setting. In the zero-shot learning setting, a model is presented with tasks or queries for which it has not received explicit training. It is expected to extrapolate knowledge from its pre-existing understanding of language and context to generate meaningful responses. This setting challenges the model to generalize effectively and showcase adaptability to novel prompts, reflecting its capacity to comprehend and manipulate language beyond the scope of its training data [21,22]. Since prompt engineering is essential when interacting with any LLM to obtain high-quality responses [23], we first experimented with and formulated prompts to elicit the desired responses from the model. Then we used our finalized prompt in zero-shot learning setting. Our finalized prompt was selected to be as follows:

> *“Task Overview:*
>
> *Given the extracted text from a research paper, identify and extract metadata related to every sensor device used in this study. The extracted information should be categorized into predefined entity labels. Ensure that the information is extracted accurately and presented in a structured JSON format:*
>
> *Entity Labels and Their Definitions:*
>
> *model_name: The term by which the instrument is known. This could be a trade name or an alias.*
>
> *model_id: The unique identifier used to differentiate each model of an instrument made by certain manufacturer.*
>
> .
>
> .
>
> .
>
> *recommended_maintenance_frequency: The frequency at which the maintenance should be repeated.*
>
> *Instructions:*
>
> - *Extract relevant metadata from the provided text file, ensuring accuracy in categorization.*
> - *If information is missing, return “N/A” for that field.*
> - *Output the extracted metadata in a JSON format. Do not include additional information. I only need the JSON structured metadata for the sensors mentioned in this paper: < text here>“.*

As a first step, we employed “*pdfplumber*” a Python library to convert each PDF document into a plain text file to ensure that the content could be effectively processed. These text files were then passed to the GPT-4o model based on the developed prompt. The model’s responses, which contained the extracted sensor metadata, were programmatically captured and saved in Excel format (Figure 1).

**Figure 1.**
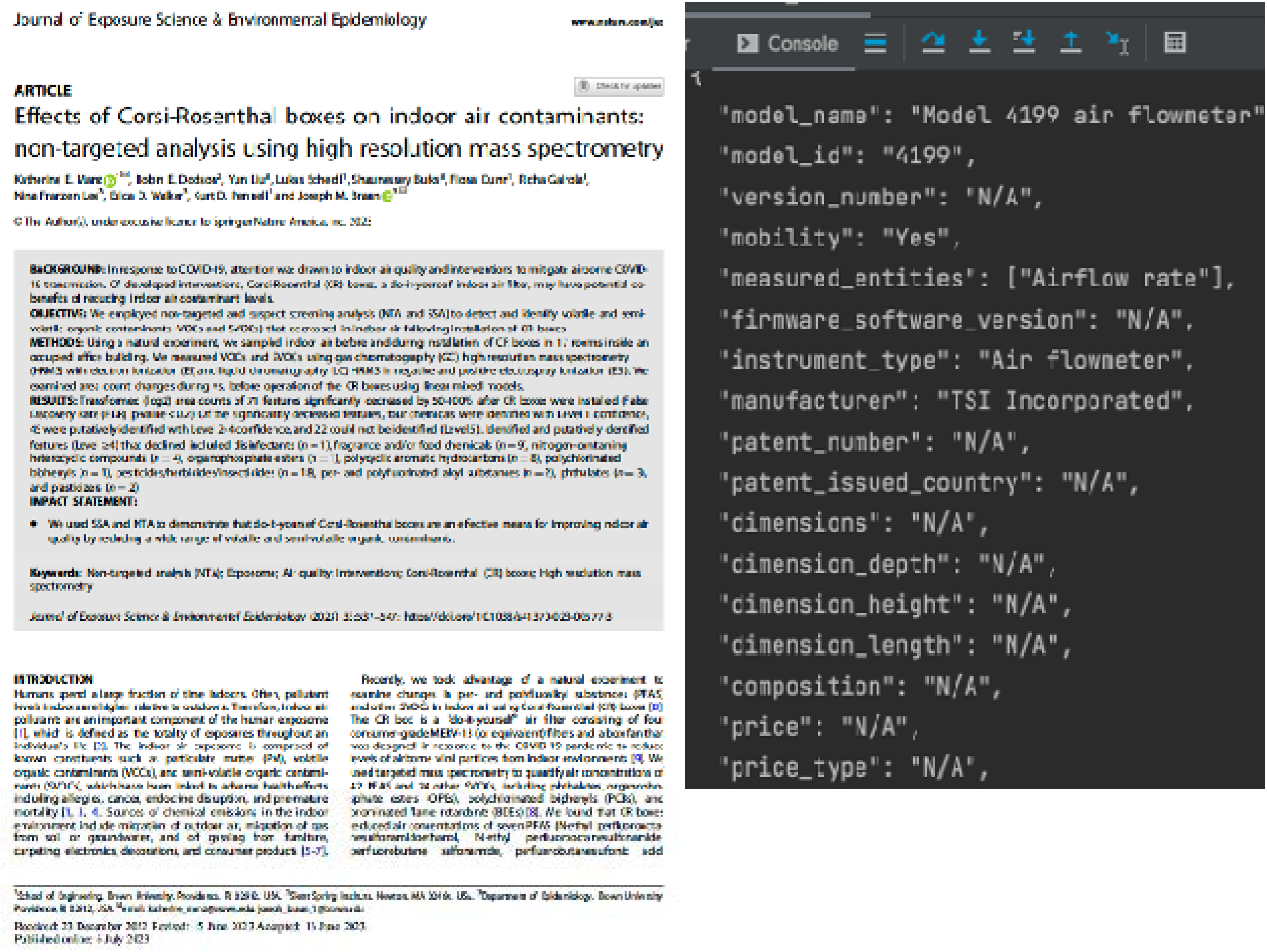
Example of input to GPT-augmented search (published journal article [34]) and output (sensor metadata in JSON format).

Despite explicitly instructing the model to return output in strict JSON format, the actual responses from GPT-4o exhibited notable variation across documents. These inconsistencies included extra quotation marks, commas, or introductory phrases such as “the first sensor metadata” preceding the structured content. As a result, the format of GPT’s output was not uniform, making automated parsing and extraction of metadata challenging. To address this, we performed a manual review of all 10 outputs (already saved in an Excel file) to identify a generalized pattern capable of isolating the JSON-formatted metadata from surrounding text. We refined a regular expression pattern that could robustly extract JSON blocks from all papers regardless of their different formatting and validated this pattern on a new (previously unused) paper to ensure its generalizability. Each block is then parsed and appended to a growing Python DataFrame. Once parsing is complete, the full set of extracted metadata is saved as an Excel file, with one file generated per paper for downstream analysis.

Following the established methods for assessing agreement [42] two human experts independently performed metadata extraction by manually reviewing the same 10 research articles and extracted the same sensor metadata attributes for all sensors reported in the papers. During this process, timestamps were also recorded to document the duration of the human review. To assess the efficiency of the automated extraction pipeline, we recorded the processing time required for each paper, from the initial PDF-to-text conversion, through GPT-based metadata extraction, to save the final structured metadata in Excel format. These timestamps were systematically logged to enable direct comparison with the time taken by human experts to manually extract metadata from the same set of documents. To quantify the performance of the GPT model against human expert annotations, we computed four standard evaluation metrics: precision, recall, accuracy, and F1-score. These metrics were calculated independently for each sensor and then aggregated to assess overall model effectiveness.

## Results

Through the manual review, a total of 15 sensors were identified across 10 research papers. Table 1 presents the results of this manual extraction, showing how frequently each metadata attribute related to the “Instrument” entity was actually reported in the articles. Each row corresponds to the number of sensors (out of 15) for which that attribute was found and could be extracted manually (Coverage n), and the percentage of total sensors with that attribute reported (Coverage %). The entities “mobility”, “measured_entities”, “instrument_type”, and “indoor_outdoor_use” were reported for 100% of sensors. “model_name”, “is_personal_device”, and “is_wearable_device” were also frequently included, appearing in over 85% of the sensors documented in the papers. In contrast, many technical or specification-level attributes such as “version_number”, “firmware_software_version”, “battery_capacity”, “output_voltage”, “charger”, and “warranty_time” had 0% coverage, indicating that these details were not reported in any of the reviewed articles. As depicted in Table 1 the coverage of “Instrument” attributes in different papers is not homogeneous.

**Table 1:** Coverage of papers for attributes for the “Instrument” Entity.

| Attribute | Coverage (n) | Coverage (%) |
| --- | --- | --- |
| Model_name | 13 | 86.67 |
| Model_id | 8 | 53.33 |
| Version_number | 0 | 0.00 |
| Mobility | 15 | 100.00 |
| Measured_entities | 15 | 100.00 |
| Firmware_software_version | 0 | 0.00 |
| Instrument_type | 15 | 100.00 |
| Manufacturer | 9 | 60.00 |
| Patent_number | 0 | 0.00 |
| Patent_issued_country | 0 | 0.00 |
| Dimensions | 2 | 13.33 |
| Dimension_depth | 0 | 0.00 |
| Dimension_height | 0 | 0.00 |
| Dimension_length | 0 | 0.00 |
| Composition | 5 | 33.33 |
| Price | 0 | 0.00 |
| Price_type | 0 | 0.00 |
| Indoor_outdoor_use | 15 | 100.00 |
| Is_personal_device | 13 | 86.67 |
| Is_wearable_device | 13 | 86.67 |
| Is_water_or_splash_proof | 0 | 0.00 |
| Needs_power_source | 8 | 53.33 |
| Power_source | 3 | 20.00 |
| Battery_operation_time_limit | 1 | 6.67 |
| Battery_capacity | 0 | 0.00 |
| Output_voltage | 0 | 0.00 |
| Is_rechargeable | 0 | 0.00 |
| Battery_type | 0 | 0.00 |
| Charger | 0 | 0.00 |
| Time_to_full_charge | 0 | 0.00 |
| Has_display | 0 | 0.00 |
| Number_of_displays | 0 | 0.00 |
| Display_type | 0 | 0.00 |
| Warranty_time | 0 | 0.00 |
| Warranty_condition | 0 | 0.00 |
| Lifetime_of_device | 0 | 0.00 |
| Recommended_maintenance_method | 1 | 6.67 |
| Recommended_maintenance_frequency | 1 | 6.67 |

### Manual Evaluation

Following the established methods for assessing agreement [42] for each of the 15 sensors and every Instrument-domain attribute, the experts created a reference (the actual value when the attribute was reported, or “N/A” when it was absent). For each attribute, our pipeline returned either a textual value or the literal string “N/A” when the paper provided no information. Both experts compared GPT’s output to the reference and labeled it agree when (i) the attribute was present and the returned value matched the reference, or (ii) the attribute was absent and GPT correctly returned “N/A.” All other cases were labeled ‘not agree’ (e.g., a wrong value for a present attribute, or a hallucinated value when the attribute was absent) along with an explanation. For each sensor we summarized these binary judgments across all attributes to compute accuracy, precision, recall, and F1: true positives were agree cases where an attribute was present and the value was correct; false negatives were not agree cases where an attribute was present but GPT returned an incorrect value or “N/A”; false positives were not agree cases where an attribute was absent but GPT returned a value; and true negatives were agree cases where an attribute was absent and GPT returned “N/A.” Table 2 presents the performance of the GPT, in terms of accuracy, precision, recall and F1-score, in comparison to the manual extraction conducted by the experts. Metrics were calculated per sensor over all attributes, and the final row reports macro-averages: average of each metric across the 15 sensors.

**Table 2:** Performance metrics per sensor and overall average.

| Sensor # | Accuracy (%) | Precision (%) | Recall (%) | F1-score (%) |
| --- | --- | --- | --- | --- |
| 1 | 100.00 | 100.00 | 100.00 | 100.00 |
| 2 | 97.37 | 97.37 | 100.00 | 98.67 |
| 3 | 94.74 | 94.74 | 100.00 | 97.30 |
| 4 | 92.11 | 92.11 | 100.00 | 95.89 |
| 5 | 92.11 | 92.11 | 100.00 | 95.89 |
| 6 | 94.74 | 94.74 | 100.00 | 97.30 |
| 7 | 94.74 | 94.74 | 100.00 | 97.30 |
| 8 | 97.37 | 97.37 | 100.00 | 98.67 |
| 9 | 94.74 | 94.74 | 100.00 | 97.30 |
| 10 | 97.37 | 97.37 | 100.00 | 98.67 |
| 11 | 97.37 | 97.37 | 100.00 | 98.67 |
| 12 | 92.11 | 92.11 | 100.00 | 95.89 |
| 13 | 92.11 | 92.11 | 100.00 | 95.89 |
| 14 | 94.74 | 94.74 | 100.00 | 97.30 |
| 15 | 89.47 | 89.47 | 100.00 | 94.44 |
| Average | 94.74 | 94.74 | 100.00 | 97.28 |

Except for one sensor, the GPT achieved accuracy and precision ranging from 92.11% to 97.37%, with recall consistently at 100% across all cases. The F1-score, which reflects the balance between precision and recall, ranged from 95.89% to 98.67%. Average accuracy and precision were 94.74%, and average recall and F1-score were 100.00% and 97.28%, respectively.

The comparison of extraction time between manual and automated methods reveals a substantial efficiency gain achieved through automation. On average, manual extraction of sensor metadata from each paper took approximately 2,000 seconds (about 33 minutes), while the automated pipeline completed the same task in just 10.7 seconds. This results in a mean time saving of 1,983 seconds per document, translating to a 99.44% reduction in execution time. Overall, the automated method was approximately 229 times faster than the manual process.

## Discussion

The GPT demonstrated the ability to infer metadata values that were not explicitly stated in the text. In multiple cases, the model successfully filled in missing details through contextual reasoning. All such inferred values were verified to be correct by expert reviewers. This suggests that GPT can effectively generalize from surrounding content to enhance metadata completeness. In some cases, the GPT extracted relevant non-sensor devices such as air pumps, treating them as distinct devices. While these instruments may not meet strict definitions of a sensor, they form integral parts of data acquisition instrumentation and collection workflows and impact the quality of the generated data. Further refinement of the prompting, either through few-shot examples that clarify the distinction between sensors and supporting devices, or by incorporating chain-of-thought reasoning can guide the model through more nuanced classification decisions.

Most articles in the corpus were primarily focused on the application of sensor devices in exposure health studies rather than on the design or engineering specifications of the sensors themselves. As a result, metadata was often sparse or incomplete both for automated and manual extraction methods. Table 2 highlights this issue. While some basic descriptive metadata was consistently available, more granular or engineering-specific metadata was often omitted. Continuing the issue of metadata sparsity, many articles referenced external sources for sensor specifications rather than describing them directly. This practice, along with the common placement of sensor details in brief “Methods” sections, often limited the extractable information about the sensors. One article described downstream analysis using chromatography but did not provide details on how the air samples were originally collected instead referencing an external publication. Future extractions could benefit from citation traversal to retrieve linked sampling meta.

Overall, the GPT-based pipeline substantially outperformed manual methods in speed of metadata extraction. It demonstrated high precision, recall, and overall performance, highlighting the strong potential GPT to streamline and scale sensor metadata extraction with minimal loss in quality. Despite its strong potential, the pipeline also revealed the need for a more adaptive and agentic approach to metadata extraction. Since metadata is often dispersed across multiple sources, an effective extraction framework must be capable of traversing citations and external references, reconciling conflicting metadata values across documents and against existing records in a metadata repository and querying the internet for supplemental information. Additionally, the pipeline should integrate human-in-the-loop mechanisms to support adjudication and expert curation, as well as track versioning and updates to stored metadata values over time.

This study demonstrates the use of AI methods to make sensors and in turn the exposome FAIR (Findable, Accessible, Interoperable, and Reusable) [40-41]. By reducing the effort required to discover and harmonize sensor metadata, this FAIRification of sensors, provides means to easily select, obtain, deploy and reuse sensor across exposure health, and provide metadata-enrichened approach to construct exposomes by assimilating exposure profiles from prospective sensor deployments as well as historic, ambient and other real-world measurements.

## Conclusion and future work

This study presents a novel application of LLMs, specifically GPT-4, to automate the extraction of sensor metadata from unstructured exposure health literature. By focusing on the “Instrument” entity within the SMARTER metadata model, we demonstrated that GPT can significantly reduce the time and labor required for metadata curation, achieving extraction speeds 229 times faster than manual review and maintaining high performance across key evaluation metrics. Our results confirm the feasibility of using LLMs for structured metadata extraction in domains where sensor documentation is often sparse, heterogeneous, and unstandardized. Moreover, the GPT showed strong contextual reasoning, successfully inferring missing values, but occasionally misclassified supporting instruments as sensors. Our findings underscore the need for future work focused on developing more adaptive pipelines that incorporate citation traversal, metadata repository integration, and human-in-the-loop curation. Enhancing these capabilities will enable scalable, accurate, and continuously updated metadata infrastructure to support sensor integration in exposure health research.

## Data Availability

All data produced in the present work are contained in the manuscript.

## Ethics Approval

No protected health information was collected or used for this work.

## Acknowledgements

This research was supported by the NIEHS, 1R24ES036134 [SMARTER], NCATS, UL1TR002538, UM1TR004409 [CTSI]. The content is solely the responsibility of the authors and does not necessarily represent the official views of the National Institutes of Health.

## Conflicts of Interest

None declared.

## Abbreviations

GPT: Generative Pre-trained Transformer
LLM: Large Language Model
SMARTER: Sensors and Metadata for Analytics and Research in Exposure Health

## Notes

### Competing Interest Statement

The authors have declared no competing interest.

### Author Declarations

This study used openly available published papers.

