## Appendix 1 for "Scaling Sensor Metadata Extraction for Exposure Health Using LLMs"

The following table lists the attributes for the entity “Instrument”, along with brief descriptions:

Table 1: Attributes for the “Instrument” Entity in the SMARTER Sensor Metadata Model [4,8]

| Attribute | Description |
| --- | --- |
| instrument_id | Auto-generated |
| model_name | The term by which the instrument is known. This could be a trade name or an alias. |
| model_id | The unique identifier used to differentiate each model of an instrument made by certain manufacturer. |
| version_number | The current version of the instrument model. It differentiates instruments within the same model. The usually refers to a version of the hardware. |
| mobility | Whether the instrument can be moved around for measuring the species. |
| measured_entities | List of all the types of measurement done by the instrument |
| firmware_software_version | Current firmware or software version of the model. |
| availability_status | Status of the Instrument - whether it is active, discontinued, or recalled etc |
| instrument_type | The category of instrument based on the species measured by the instrument. |
| manufacturer | The person, group, or organization that develops or produces the instrument. |
| patent_number | The serial number of the patent, if the instrument is patented. |
| patent_issued_country | The country issuing the patent. |
| dimensions | The size of the instrument in physical space. The dimension could have attributes of depth, height, and length. Each dimension includes a value and unit. (Use this if the dimensions aren’t available discretely, else use the below fields.) |
| dimension_depth | The depth or thickness of the instrument. |
| dimension_height | The vertical height of the instrument. |
| dimension_length | The horizontal length of the instrument. |
| composition | The description of the composition of combining parts or elements making up of the instrument. |
| price | The cost of the instrument. This could be a potential price or price range of the instrument, such as the manufacturer recommended price, actual price, or price range to purchase the instrument. |
| price_type | Whether the price/price range is the potential price or actual price to purchase the instrument. |
| indoor_outdoor_use | Whether the instrument is intended to be used inside a building or structure that is protected from the natural environment. Or, if the instrument can be used outdoors and can tolerate exposure to the natural environment. |
| is_personal_device | Whether or not the instrument is intended to be used to and track information for individuals. |
| is_wearable_device | Whether or not the instrument can be worn by individuals on their body or carried, and track information. |
| is_water_or_splash_proof | Whether or not the instrument can tolerate exposure to water. |
| needs_power_source | Whether or not the instrument needs a source of power for its normal function. If power is needed, the type of power should be listed. See "Source of Power." |
| power_source | The type of power that supports the instrument for its normal function/s. |
| battery_operation_time_limit | The duration of battery life, if the "source of power" is battery. |
| battery_capacity | The amount of electric charge the battery can deliver at the rated voltage. |
| output_voltage | The voltage released by the battery. |
| is_rechargeable | Whether or not the battery's electric charge can be restored by connecting the battery to a recharging device. |
| battery_type | The category of battery, based on the chemical used in the battery's electrochemical cells. |
| charger | If the battery is rechargeable, this element is used to describe the charger. |
| time_to_full_charge | The time taken to recharge the battery. |
| has_display | Whether or not the instrument is capable of displaying information. If yes, more information can be recorded in the following data element, such as how many monitors, and what type of monitors does it have. |
| number_of_displays | The number of displays with the instrument. |
| display_type | The category of the monitor used to display information. |
| warranty_time | The length of time covered by the instrument's warranty. |
| warranty_condition | The facts or conditions under which the warranty is valid. |
| lifetime_of_device | The duration of time during which the instrument is expected to function properly according to the manufacturer. |
| recommended_maintenance_method | The method suggested for maintaining the instrument. |
| recommended_maintenance_frequency | The frequency at which the maintenance should be repeated. |
