## Supplementary material for "Scaling Sensor Metadata Extraction for Exposure Health Using LLMs": Figure 1

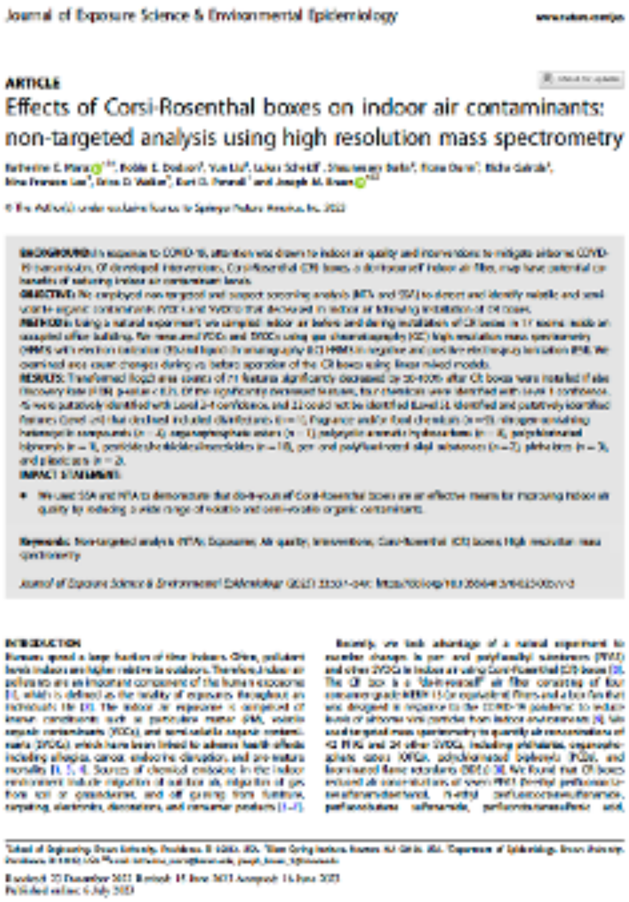
Journal of Exposure Science & Environmental Epidemiology

ARTICLE

### Effects of Corsi-Rosenthal boxes on indoor air contaminants: non-targeted analysis using high resolution mass spectrometry

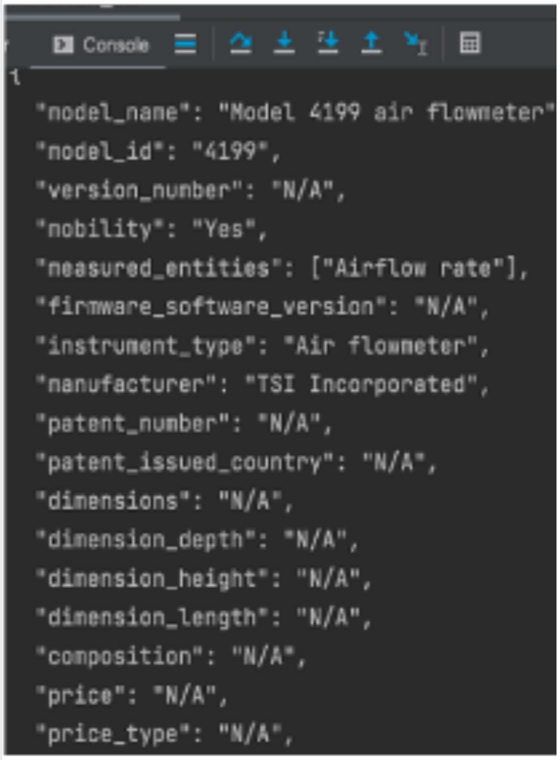



```
{
  "model_name": "Model 4199 air flowmeter"
  "model_id": "4199",
  "version_number": "N/A",
  "mobility": "Yes",
  "measured_entities": ["Airflow rate"],
  "firmware_software_version": "N/A",
  "instrument_type": "Air flowmeter",
  "manufacturer": "TSI Incorporated",
  "patent_number": "N/A",
  "patent_issued_country": "N/A",
  "dimensions": "N/A",
  "dimension_depth": "N/A",
  "dimension_height": "N/A",
  "dimension_length": "N/A",
  "composition": "N/A",
  "price": "N/A",
  "price_type": "N/A",
```
